# Immunogenomic profiling of lung adenocarcinoma reveals high-grade growth patterns are associated with an immunogenic tumor microenvironment

**DOI:** 10.1101/2022.03.17.22272385

**Authors:** Neal Akhave, Erin M. Bayley, Meredith Frank, Carmen Behrens, Jiexin Zhang, Runzhe Chen, Xin Hu, Edwin Roger Parra, Won-Chul Lee, Stephen Swisher, Luisa Solis, Annikka Weissferdt, Cesar Moran, Neda Kalhor, Jianhua Zhang, Paul Scheet, Ara A. Vaporciyan, Boris Sepesi, Don L. Gibbons, John V. Heymach, Jack J. Lee, Ignacio I. Wistuba, P. Andrew Futreal, Jianjun Zhang, Junya Fujimoto, Alexandre Reuben

## Abstract

Lung cancer is the leading cause of cancer-related mortality in the United States. Lung adenocarcinoma (LUAD) is the most common subtype and the most epidemiologically and genetically heterogeneous. Pathologists have routinely observed phenotypic heterogeneity among LUAD primary tumors as reflected by distinct patterns of tumor growth. However, despite prior implications on the association of immune-genomic environment and prognosis, this information is not utilized clinically. Herein, applying multiplatform immune-genomic analysis, we investigate two distinct classification systems and demonstrate that high-grade patterns of growth are associated with a distinct immunogenic tumor microenvironment that is predicted with a favorable response to immunotherapy, a finding with growing importance in the era of adjuvant and neoadjuvant immunotherapy.

## Introduction

Lung cancer remains the leading cause of cancer-related mortality in men and women in the United States^1,2^. However, over the course of the last century, and particularly with the advent of genomic sequencing, there have been significant advances in the understanding of lung cancer risk factors, genetic basis, and treatments. Small cell and non-small cell lung cancer (NSCLC) are regarded as the most clinically meaningful subtypes of lung cancer^3,4^ with NSCLC further sub-classified as lung adenocarcinoma (LUAD) and squamous cell carcinoma (LUSC) due to distinct genetic basis and disparate responses to systemic therapy^5-7^.

LUAD is the most common subtype and the most epidemiologically and genetically heterogeneous^5^. Tobacco exposure is the most prevalent risk factor for LUAD; however, nearly 20-30% of cases are identified in never smokers^2^. Further, LUAD in smokers is enriched for activating mutations in *KRAS*, high tumor mutation burden, and increased T-cell infiltration and clonality^5,8^. In contrast, LUAD in never smokers is rarely associated with *KRAS* alterations and predicts for the presence of a diverse set of oncogenic mutations or fusions, each with distinct disease biology and treatment^3,5,8^.

However, pathologists have routinely observed phenotypic heterogeneity among LUAD primary tumors as reflected by distinct patterns of tumor growth which prompted the International Association for the Study of Lung Cancer (IASLC)/American Thoracic Society (ATS)/European Respiratory Society (ERS)^9^ to develop a multidisciplinary classification that was subsequently incorporated into the World Health Organization (WHO) classification of LUAD^10^. This classification system identified five distinct patterns of growth in LUAD: acinar, lepidic, papillary, micropapillary, and solid^11^, which were classified by predominant subtype.

Multiple clinical studies have attempted to determine the relationship of patterns of growth and genomic driver of prognosis and have suggested that solid or micropapillary tumors are high-grade and associated with worse prognosis^12-22^. However, due to large-scale differences in sample size and clinical variables such as completeness of surgical resection or pathologic stage, findings have not been uniformly consistent. Thus, at this time, LUAD patterns of growth have had limited use in clinical decision-making. However, particularly in the immunotherapy and cellular therapy era, further study is warranted to understand the relationship of LUAD pattern of growth and immune activation and senescence within the tumor microenvironment that could affect efficacy of these novel agents.

Herein, we describe the most comprehensive examination of the relationship between LUAD pattern of growth with genomic-immunologic disease correlates and survival. Using the PROSPECT (Profiling of Resistance patterns and Oncogenic Signaling Pathways in Evaluation of Cancers of the Thorax; LAB07-0233) patient cohort from the University of Texas M.D. Anderson Cancer Center, a robust dataset of stage I-III NSCLC tumors collected from 1996 to 2008, we applied two distinct classification methodologies to understand the relationship between LUAD patterns of growth and their relationship with baseline clinical variables, prognosis, and immunogenomic environment.

Our study reaffirms conclusions of prior studies that high-grade patterns of growth are associated with worse relapse-free and overall survival; however, in contrast to prior studies, this relationship is the result of an association of high-grade patterns with higher stage of disease at time of surgery. Additionally, we demonstrate that when tumors are reclassified by a risk-based tiered fashion, the presence of a high-grade pattern of growth of any percentage is associated with prolonged tobacco exposure, elevated tumor mutation burden, and an immune infiltrated tumor microenvironment with increased cytotoxic CD8 T-cell infiltration, PD-L1 expression, and decreased T-cell repertoire homology with tumor-adjacent lung tissue, which may predict for differences in therapeutic response to novel immunotherapies.

## Methods

### Patient Cohort and Sample Collection

After informed consent, enrollment of study participants and collection of study samples was performed via the PROSPECT (Profiling of Resistance patterns and Oncogenic Signaling Pathways in Evaluation of Cancers of the Thorax; LAB07-0233) study approved by the University of Texas M.D. Anderson Cancer Center’s Institutional Review Board (IRB). Between 1996 and 2008, fresh-frozen and formalin-fixed paraffin-embedded (FFPE), peripheral blood mononuclear cells (PBMC), serum, tumor, and tumor-adjacent lung tissue samples were obtained from 174 LUAD patients at time of diagnosis and surgery. Histological subtype was defined and pathologic classification was performed on all samples and patients were treatment-naïve at the time of surgery. As part of pathologic sub-classification, patterns of growth were quantified within tumors such that the sum total of all patterns within an individual tumor was 100%. LUAD tumors were classified using two distinct classification systems: a predominant-pattern method, as per IASLC-ATS-ERS and WHO, and a risk-based tiered method. In the predominant-pattern method, LUAD tumors were classified by the largest single pattern of growth and invasion (*i*.*e* acinar, lepidic, papillary, or solid) as defined by relative percentage. Conversely, in the risk-based tiered method, tumors were simply classified by the presence or absence of high- (*i*.*e*. solid) or low-grade (*i*.*e*. lepidic) patterns of growth, independent of relative percent. Tumors with papillary and/or acinar patterns of growth were classified as intermediate-grade and were classified by the predominant pattern. For the purposes of comparison, independent of classification method, solid tumors were designated as high-grade LUAD group, and all lepidic, papillary, and acinar tumors were combined to form the low/intermediate-grade LUAD group.

### TCR variable beta chain sequencing

Sequencing of the CDR3 regions of the human TCR-β chains was performed using the immunoSEQ Assay (Adaptive Biotechnologies, Seattle, WA)^23-25^. T cell density, richness, clonality, and similarity were calculated as previously described^8,26^. Briefly, TCRβ counts were normalized to the total amount of DNA usable for TCR sequencing as determined by PCR-amplification and sequencing of housekeeping genes before calculating T cell density. The preseqR package was used to calculated richness by extrapolating to 400,000 templates for PBMCs and 120,000 templates for tissue. To allow comparison of samples with unequal numbers of T cells, both richness and clonality normalize for sampling depth. Clonality was defined as 1-Peilou’s eveness^27^. As described previously, differential abundance analyses were used to identify TCRs that were enriched in one sample over another^28^. Parameters were as follows: minTotal = 5, productiveOnly = True, alpha = 0.1, count = aminoAcid. Statistical analysis was performed in R version 3.2.

### Whole Exome Sequencing

Whole exome sequencing (WES) was performed on tumors and spatially segregated tumor-adjacent lung tissues to determine somatic point mutations in a prior study^29,30^, where single nucleotide variants (SNV) and small insertions and deletions (indels) were detected using MuTect^31^ and Pindel^32^, respectively., followed by variants annotation and filter^33^. In addition, genomic DNA from 96 available matched peripheral blood samples was sequenced as germline control to identify the mutations in the tumor-adjacent lung tissues. Blood DNA was analyzed to identify mutations related to clonal hematopoiesis of indeterminate potential (CHIP) based on annotation specified previously^34^. WES data is available in the EGA (EGAS00001004026).

### Gene Expression Profiling

RNA microarray was performed in a prior study on 141 patients^35,36^ using the Illumina HumanWG-6 v3.0 expression bead chip. Then an extended robust multi-array analysis (RMA) background correction model^37^ was applied to obtain normalized gene expression profiles for individual samples. Gene expression data is available in the GEO repository (GSE42127).

### CIBERSORT

To estimate the abundances of different cell types in mixed cell population, we ran CIBERSORT ^38^ (https://cibersort.stanford.edu) using leukocyte gene signature matrix (LM22) containing 22 functionally defined human immune subsets to deconvolute our normalized gene expression profiles with default settings.

### Immunohistochemistry

Tumor tissue was fixed in formalin and embedded in paraffin. For immunohistochemical staining, tissue was cut and mounted at a thickness of 4μm per slide. Slides were then stained with CD3 polyclonal (1:100, DAKO), CD4 clone 4B12 (1:80, Leica Biosystems), CD8 clone C8/144B (1:25, Thermo Scientific), PD-L1 clone E1L3N (1:100, Cell Signaling Technology), PD-1 clone EPR4877-2 (1:250, Abcam), CD45RO clone UCHL1 (ready-to-use, Leica Biosystems), FoxP3 clone 206D (1:50, BioLegend), and Granzyme B clone F1 (ready-to-use, Leica Biosystems)^39^ antibodies. Slides were then stained using diaminobenzidine as chromogen and the Leica Bond Polymer refine detection kit (Leica Biosystems). Slides were then counterstained with hematoxylin and scanned using an Aperio AT2 automated slide scanner (Leica Biosystems). Quantification was performed on 5 × 1mm2 regions per tumor sample within the tumor center and measuring the average density of positive cells per region as a count of positive cells/mm2. For PD-L1, H-score was calculated by multiplying the proportion of positive cells in the sample (0– 100%) by the intensity of staining (1+, 2+, or 3+) to obtain a score ranging between 1 and 300.

### Statistical Analysis

All plots were generated using GraphPad Prism 8.0 (La Jolla, CA). For baseline characteristics, for continuous variables, t-test was applied. Further, for categorical variables, based upon sample size of < or >5, chi-square or Fisher’s exact test was applied. Because not all distributions of TCR variables met the normality assumption, Mann-Whitney U test or Kruskal-Wallis test (two-sided) was applied for assessing differences among groups. Wilcoxon signed-rank test was used to compare matched samples. Spearman’s rank correlation (two-sided) was used to assess monotonic relationships between two continuous variables. For survival analysis, we first performed Log-rank (Mantel-Cox) test. Finally, despite the exploratory nature of the study, p-values were adjusted for multiple hypothesis testing (p-value < 0.05).

## Results

### Risk-based tiered system reclassifies tumors by presence of any amount of high-grade pattern of growth and demonstrates relationship with smoking and stage

One hundred and seventy-four patients with LUAD enrolled onto the PROSPECT study were included in our analysis. All patients underwent curative-intent surgery followed by classification of histologic pattern of growth by expert thoracic pathologist and comprehensive multi-platform profiling. Dual classification systems were utilized to ensure that the commonly used predominant-pattern (IASLC/ATS/ERS) methodology would not disregard smaller percentage tumor patterns, particularly high-grade growth patterns, whose molecular contribution to the tumor microenvironment would be masked within the context of an otherwise-classified low/intermediate grade tumor. Thus, we devised a risk-based tiered classification method to stratify tumors for the presence or absence of high-risk tumor components and then assessed relationship with tumor microenvironment. Finally, before comparative analysis, independent of classification method, solid tumors were placed in the high-grade group and lepidic, papillary, and acinar tumors were combined to form the low/intermediate grade group (**Supplementary Figure 1a-b**). With this, as shown in **Supplementary Figure 1b-c**, when comparing the predominant pattern to risk-based tiered classification method, 40 (23%) LUAD tumors classified as low/intermediate grade by predominant pattern classification (i.e. containing ≤ 49% solid pattern of growth) were reclassified as high-grade using the risk-based tiered classification.

Comparison of baseline characteristics, as shown in **Supplementary Table 1** (predominant pattern classification) and **Supplementary Table 2** (risk-based tiered classification), identified high-grade tumors as enriched for association with active and heavy tobacco use, as defined by current smoking and smoking for >20 pack-years, respectively. Notably, differences were particularly pronounced when utilizing the risk-based tiered method (**Supplementary Table 2**) with 54% and 80% of high-grade group patients being either current active smokers or having >20 pack-year history of smoking, relative to only 26% (*p=0*.*001*) or 58% (*p=0*.*002*) of low-intermediate grade group patients.

Further, comparison of pathologic stage using both classification systems (**Supplementary Table 1; Supplementary Table 2**) demonstrated a significant association of high-grade tumors with advanced stage by the predominant pattern classification with 64% of high-grade tumors being stage 2 or 3, in contrast to only 37% of low/intermediate grade tumors (*p=0*.*001)*. However, this difference was not observed when using the risk-based tiered classification (*p=0*.*59*). Of note, as shown in **Figure 1a-c**, no association was observed between percentage of high-grade growth pattern and primary tumor size (T-stage) (**Figure 1b**; r=0.18; p=0.08; Spearman rank correlation). However, increasing percentage of high-grade growth pattern was strongly associated with presence of nodal metastases (N-stage) (**Figure 1c**; p<0.001). Altogether, these findings suggest that when high-grade patterns of growth are dominant, disease is likely of advanced stage with increased risk of nodal metastases, which may reflect a more invasive tumor biology and warrants evaluation about the need for more thorough lymph node evaluation pre-or intra-operatively.

**Figure 1:**
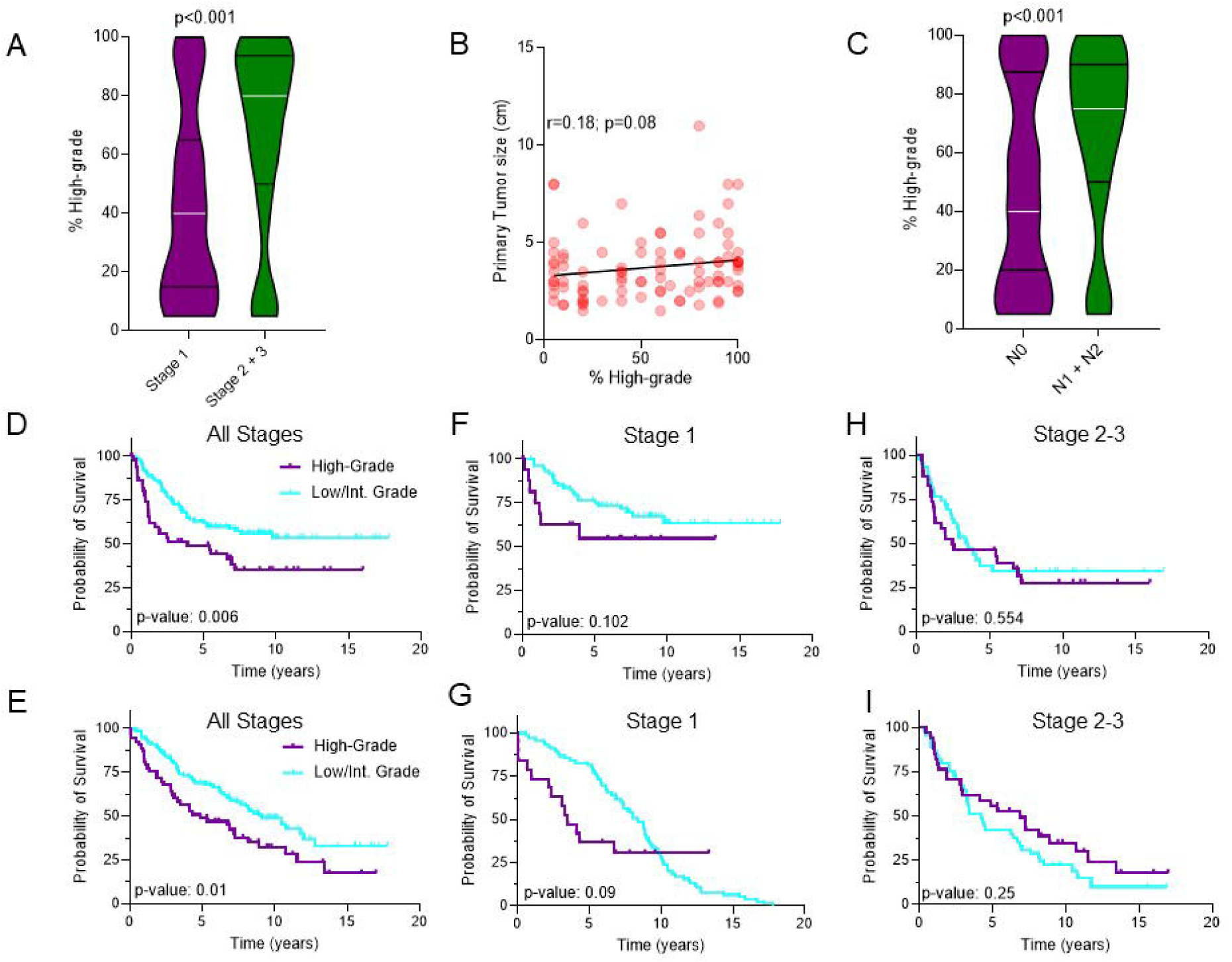

### High-grade pattern, when predominant, *may* be associated with worsened disease-free & overall survival

Next, we examined the relationship between LUAD pattern of growth and prognosis. As in prior studies^12-14^, we found that the presence of high-grade growth patterns, when defined by predominant pattern, was associated with worse relapse-free (RFS) and overall survival (OS) (**Figure 1d; Figure 1e**). However, unlike prior studies which suggested that this difference was independent of pathologic stage, our findings were tied to pathologic stage with loss of survival disadvantage when stratified by stage 1 or stage 2 and 3 cohorts (**Figure 1f-i**). Nonetheless, in the stage 1 population, although not statistically significant, there was a trend towards worsened RFS and OS (**Figure 1f-g**), as has been shown previously^17,19,40^ and supports the use of the predominant pattern (IASLC-ATS-ERS) classification as the preferred method to stratify tumors to understand disease prognosis.

Use of the risk-based tiered method to assess RFS and OS (**Supplementary Figure 2a-f**) was not associated with differences in RFS or OS, even among stage 1 adenocarcinomas. This finding suggests that the impact of high-grade patterns of growth upon prognosis is present only when it is the predominant component, at least in the pre-immunotherapy era.

### High-grade pattern is associated with increased tumor mutation burden

Due to the burden of smokers in the high-grade group, we next assessed the relationship of pattern of growth and tumor mutation burden (TMB), which has been recurrently shown to be increased in lung cancer, particularly smoking-associated lung cancer, relative to other cancer types41,42. Additionally, TMB has been shown to be correlated with increased T-cell clonality and T-cell activation^8^. Finally, TMB of greater than 10 mutations per megabase has been shown to be an independent predictor of response to immunotherapy across human cancer^43^, including LUAD^44^.

First, to validate findings of prior studies in our dataset, we assessed the relationship of TMB, tobacco use and T-cell clonality. TMB demonstrated a significant correlation with pack-years of smoking (**Figure 2a;** *r=0*.*37; p=0*.*0001; Spearman rank correlation*) and T-cell clonality (**Figure 2b;** *r=0*.*28; p=0*.*02*; *Spearman rank correlation*). Next, we compared TMB between high-grade and low-intermediate grade tumors by total non-synonymous exonic mutations (NSEM) and by number of tumors with >10 mutations/megabase (Mut/Mb) or <10 Mut/Mb. Utilizing the predominant-pattern classification, high-grade tumors showed no association with TMB by total NSEM (**Supplementary Figure 3a;** *p=0*.*52*). However, high-grade tumors were significantly enriched for tumors with >10 Mut/Mb (**Supplementary Figure 3b;** *p=0*.*02*). Further, when comparing TMB by risk-based tiered methodology, differences in TMB were even more pronounced with high-grade tumors showing significant increase in TMB relative to low/intermediate grade tumors, both in total NSEM and number of tumors with >10 Mut/Mb. Specifically, high-grade tumors were found to have 273 (ranging from 8 to 857) NSEM per tumor compared to only 116 (ranging from 3 to 653) NSEM per tumor in low-intermediate grade tumors (**Supplementary Figure 4a;** *p=0*.*046*). Additionally, high-grade tumors showed significant enrichment for tumors with >10 Mut/Mb (**Figure 2c;** *p=0*.*008*) with 12 (86%) of tumors with >10 Mut/Mb identified as high-grade.

**Figure 2:**
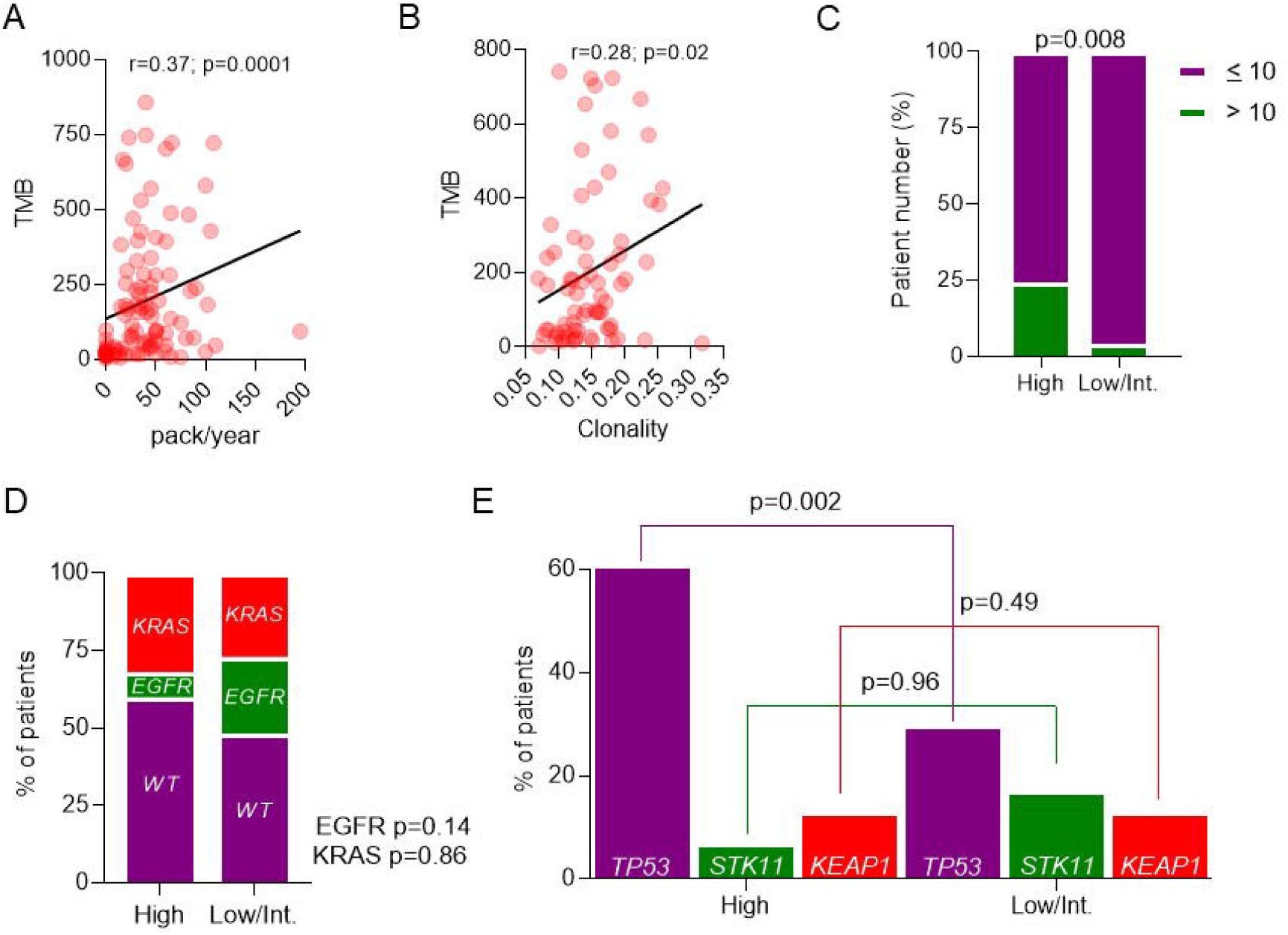

Altogether, these findings suggest that high-grade patterns, as identified by both classification systems, are associated with increased TMB. However, the risk-based tiered classification system more clearly stratifies tumors by TMB and suggests that high-grade pattern at any percentage within a tumor may predict for increased responsiveness to immunotherapy, in part due to its association with increased TMB.

### High-grade pattern is associated with mutations in *TP53*

We next assessed the relationship between high and low/intermediate grade pattern and known LUAD oncogenic drivers, *KRAS* and *EGFR*, and tumor suppressor genes, *TP53, STK11*, and *KEAP145*. Notably, although whole exome sequencing was used in this dataset, fusions were poorly captured and therefore we were unable to assess for presence of fusion-specific oncogenic drivers in LUAD involving *ALK, ROS1*, and *RET* genes. Neither predominant-pattern classification nor risk-based tiered method showed an enrichment of activating mutations in *KRAS* or *EGFR* (**Supplementary Figure 3c; Figure 2d**) or loss of function mutations in *STK11 or KEAP1* (**Supplementary Figure 3d; Figure 2e**). However, both classification methods demonstrated significant enrichment for mutations in *TP53* (**Figure 2d;** *p=0*.*002;* **Supplementary Figure 3d;** *p=0*.*003*). As with TMB, *TP53* mutations have been suggested to also predict responsiveness to checkpoint inhibitors,^46,47^ and create for an environment that allows for cancer cell proliferation^48^ and future genomic doubling, a finding associated with poor prognosis^48^.

### High-grade pattern is associated with increased immune infiltration and T-cell activation

As prior studies have suggested relationship of *TP53* mutations and increased TMB with increased infiltration of cytotoxic and activated CD8 T-cells and M0/M1 macrophages^49^, we next assessed the relationship between high- and low/intermediate grade tumors and tumor microenvironment. As our group has previously published upon the immune characteristics of this patient cohort using TCRseq, IHC, and GEP^8,26,29,30,35,36,39^, we reanalyzed the data to interrogate the composition of the high-grade and low/intermediate grade tumors and associated tumor-adjacent uninvolved lungs (TAL).

First, when utilizing the predominant-pattern classification, IHC revealed no differences in the density of T-cell populations within the tumor (**Supplementary Figure 5a**). However, the ratio of CD4 to CD8 T-cells was significantly elevated in low/intermediate grade tumors relative to high-grade tumors with ratio of 1.71 (ranging from 0.34 to 5.18) and 1.32 (ranging from 0.42 to 4.04), respectively (**Supplementary Figure 5b**; *p=0*.*01*). Moreover, using CIBERSORT, intra-tumoral CD4 T-cells were more likely to be of activated memory CD4 T-cell programming (**Supplementary Figure 6b;** *p=0*.*003*). However, no differences were found among other cell types (**Supplementary Figure 6a-d**).

In the TAL, high-grade tumors were notable for significant increase in density of CD4 and CD8 T-cells and Granzyme B (GZMB) relative to low/intermediate-grade tumors (**Supplementary Figure 5c**). Further, although there was no difference in CD4:CD8 ratio within TAL (**Supplementary Figure 5b**; *p=0*.*17*), the ratio of CD4 T-cells between tumor and TAL was significantly increased in low/intermediate grade tumors relative to high-grade tumors with Tumor:TAL-CD4 ratio of 0.96 (ranging from 0.22 to 2.31) and 0.78 (ranging from 0.11 to 2.0), respectively (**Supplementary Figure 5d;** *p=0*.*002*). Altogether, our findings suggest that high-grade patterns may drive a more active T-cell population both within LUAD tumors and in the tumor-adjacent lung.

Reclassification of tumors by risk-based tiered methodology was able to identify additional differences in immune composition between high-grade and low/intermediate grade tumors. Firstly, IHC was now able to identify a significantly increased density of infiltrating CD8 T-cells (*p=0*.*02)* and GZMB-expressing cells (*p=0*.*01)* (**Figure 3a**) in high-grade tumors, a difference not found with other T-cell populations (**Figure 3a; Supplementary Figure 7a**). Further, although all T-cell markers were positively correlated with GZMB, among high-grade tumors the density of GZMB was most positively correlated with CD8 T-cells (**Figure 3b;** *p=<0*.*0001; r=0*.*67; Spearman rank correlation*) as opposed to CD4 T-cells (**Supplementary Figure 7b**; *p=<0*.*0001; r=0*.*35; Spearman rank correlation*), highlighting the cytotoxic nature of infiltrating CD8 T-cells.

**Figure 3:**
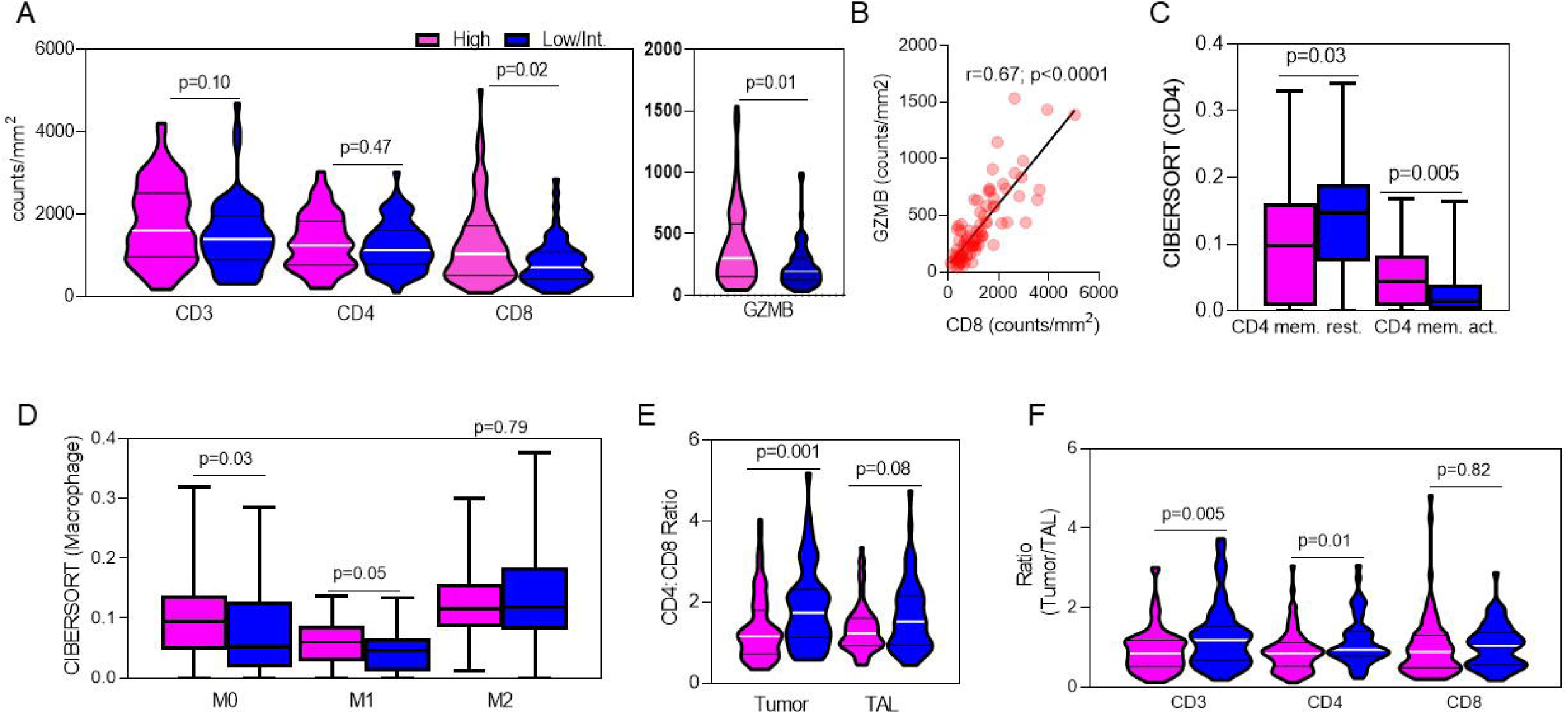

Further, functional assessment by CIBERSORT revealed that CD4 T-cells within low/intermediate grade tumors favored a resting CD4 memory cell phenotype (**Figure 3c;** *p=0*.*03*), whereas CD4 T-cells within high-grade tumors favored an activated CD4 memory cell phenotype (**Figure 3c;** *p=0*.*005*). Additionally, although no differences among nearly all other adaptive and innate immune cells were observed (**Supplementary Figure 8a-b**), high-grade tumors demonstrated slight increase in prevalence of non-activated (M0) (**Figure 3d;** *p=0*.*03*) and pro-inflammatory (M1) macrophages (**Figure 3d;** *p=0*.*05*), but not anti-inflammatory (M2) macrophages (**Figure 3d;** *p=0*.*79*). This finding suggests that high-grade tumors may be more primed to foster an anti-tumor immune response^50^ and supports the idea of a distinct immunogenic TME phenotype within high-grade tumors. Finally, the lack of increase in activated NK cell programming among high-grade tumors further supports that the observed increase in granzyme B expression is likely solely due to increased CD8 T cell infiltration (**Supplementary Figure 8b**).

Further comparison of intra-tumoral CD4:CD8 ratio revealed a further intensification of differences among high-grade and low-intermediate grade tumors. Specifically, among low/intermediate grade tumors, intra-tumoral CD4:CD8 ratio was 1.88 (ranging from 0.58 to 5.18) compared to high-grade tumors with CD4:CD8 ratio of 1.37 (ranging from 0.34 to 4.04) (**Figure 3e;** *p=0*.*001*), again suggesting enrichment of CD8+ T cells within high-grade tumors, something not seen in the TAL **(Figure 3e;** *p=0*.*08*).

Finally, review of T-cell populations within TAL revealed significantly increased T-cell density across all markers tested among high-grade tumors (**Supplementary Figure 8c**). However, notably, ratio of Tumor:TAL CD3 (*p=0*.*005*) and CD4 (*p=0*.*01*), but not CD8 (*p=0*.*82*), was higher among low-intermediate grade tumors suggesting that CD8 T cells may be less capable to infiltrate or excluded from these tumors (**Figure 3f**).

### High-grade pattern is associated with increased expression of immune checkpoints, particularly PD-L1

Considering the increased T-cell infiltration seen in high-grade tumors, we next assessed the expression of checkpoints used by malignant tumors, including LUAD, to induce immune senescence.

First, using the predominant-pattern classification, we compared PD-L1 H-score between high-grade and low/intermediate grade tumors. As shown in **Supplementary Figure 9a**, PD-L1 H-score was significantly elevated in high-grade tumors (*p=0*.*008*), however, gene expression profiling of other immune checkpoints, CTLA-4, TIGIT, and LAG-3 did not demonstrate significant differences (**Supplementary Figure 9b**).

However, analysis using risk-based tiered classification revealed not only significant enrichment of PD-L1 intensity among high-grade relative to low/intermediate grade tumors (**Figure 4a;** *p=0*.*032*) but also increase in LAG-3 expression, an immune checkpoint molecule for which checkpoint inhibitors are in development for NSCLC and already showing success in combination with PD-1 inhibitors in metastatic melanoma^51^ (**Figure 4b;** p=0.04). Altogether, these findings reinforce that even small percentages of high-grade pattern, as defined by risk-based tiered classification, unveil key differences in immunogenic potential of LUAD tumors, which may have clinical implications.

**Figure 4:**
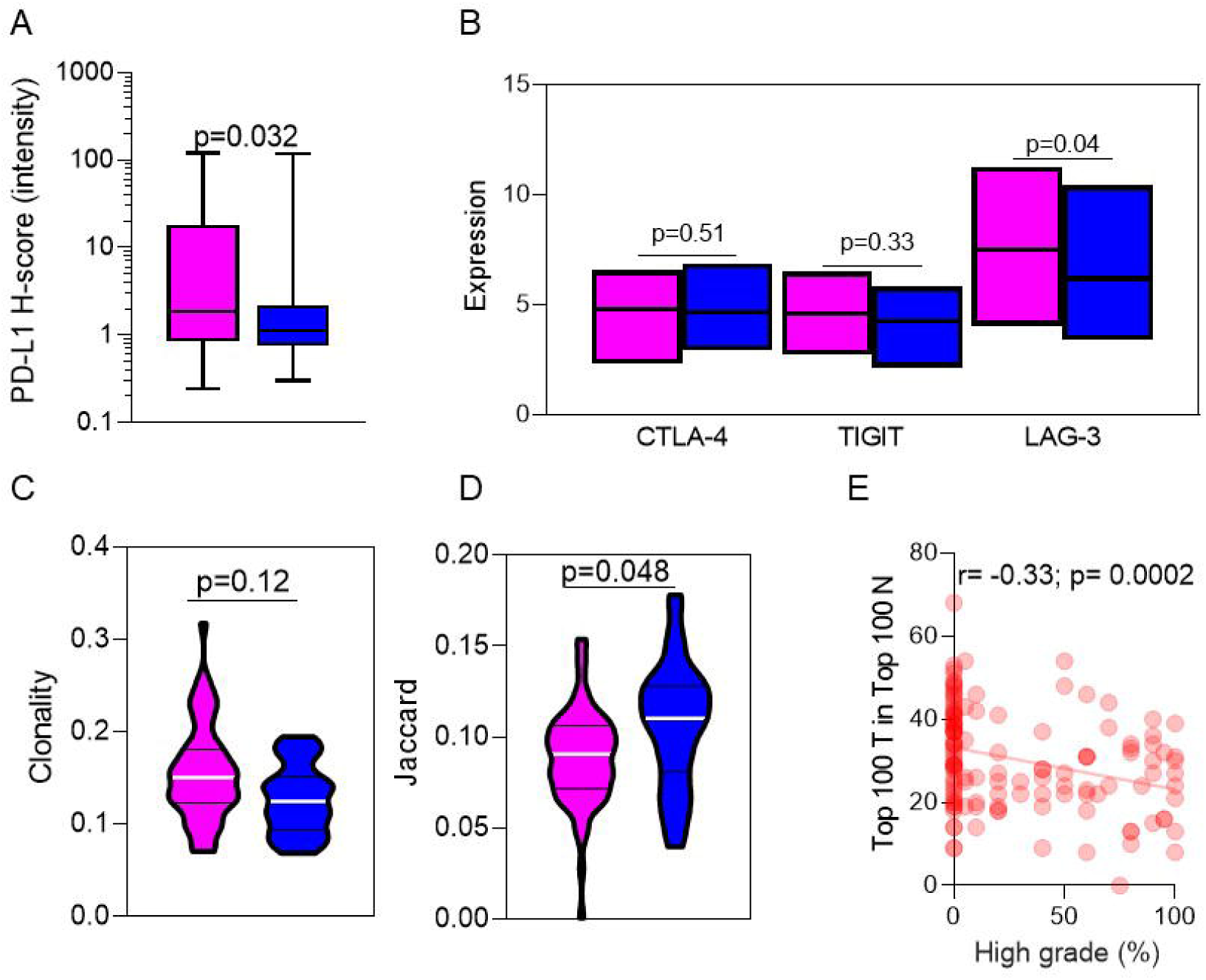

### High-grade pattern is associated with decreased T-cell repertoire homology compared to TAL

Finally, to further investigate the nature of the infiltrating T-cell repertoire between high-grade and low/intermediate grade tumors, we analyzed TCR sequencing data generated from tumor and TAL repertoires.

First, when applying the predominant-pattern classification, we identified no differences in T-cell clonality between high-grade and low-intermediate grade tumors (**Supplementary Figure 10a;** *p=0*.*10*). Further, comparison of homology between tumor and TAL by Jaccard (**Supplementary Figure 10b;** *p=0*.*58*) and Morisita indices (**Supplementary Figure 10c;** *p=0*.*30*), and total number of shared T-cell clones among top 100 clones (**Supplementary Figure 10d;** *p=0*.*27*) showed no differences between high-grade and low/intermediate grade tumors.

However, utilization of risk-based tiered classification, although again not demonstrating a difference in T-cell clonality between high-grade and low-intermediate grade tumors (**Figure 4c;** *p=0*.*12*), did show distinct differences in T-cell repertoire homology between tumor and TAL. Specifically, evaluation by Jaccard index revealed significantly greater homology of T-cell clones between tumor and TAL in low-intermediate grade tumors relative to high-grade tumors (**Figure 4d;** *p=0*.*048*), a finding that approached but did not reach significance when using the Morisita Index (**Supplementary Figure 11a;** *p=0*.*051*), suggesting differences in clonal expansion and distribution between low/intermediate and high-grade tumors. Furthermore, among the top 100 most prevalent T-cell clones identified in individual tumors, low/intermediate grade tumors had an average of 34 among the top 100 in the TAL (**Supplementary Figure 11b**). In contrast, high-grade tumors had an average of only 27 among the top 100 in the TAL (**Supplementary Figure 11b;** *p=0*.*028*). Additionally, a negative correlation was seen between increasing high-grade pattern and shared Top 100 T-cell clones (**Figure 4e;** *r=-0*.*33; p=0*.*0002; Spearman rank correlation*). Shared T-cell repertoire homology has been suggested to be non-tumor specific by a recent publication by our group^8^. Therefore, results suggest that the T-cell repertoire in high-grade tumors, as identified by risk-based tiered classification, may reflect more tumor-specific clonal expansion that may further predict and augment immunotherapy responses.

## Discussion

This study represents the most comprehensive analysis of the genomic and immune underpinnings of LUAD morphologic patterns of growth and invasion. Utilizing two distinct methods of classification, the predominant-pattern (WHO) classification and risk-based tiered classification, our study highlights the key clinical, prognostic, genomic, and immunologic differences between tumors with high-grade and low/intermediate grade patterns of growth.

We demonstrate that, when classified by the predominant-pattern method, high-grade tumors are associated with worsened relapse-free and overall survival. However, unlike prior studies which suggest that pattern of growth is an independent predictor of survival, we demonstrate that prognosis is more closely linked to pathological stage rather than pattern of growth. Further, our findings suggest that increasing percentage of high-grade growth patterns within LUAD primary tumors are associated with increased risks of nodal metastases, suggesting a more invasive phenotype among high-grade tumors. Current standard of care for mediastinal lymph node dissection is lymphadenectomy of stations 2R, 4R, 7, 8, and 9 for right-sided tumors and stations 4L, 5, 6, 7, 8, 9 for left-sided tumors^52^. However, our results suggest that in tumors with high-grade pattern of growth, even in small percentage, expanded lymph node dissection to capture occult metastases may be necessary.

Further, we suggest that the predominant-pattern classification is ill equipped to identify meaningful differences in immune microenvironment of LUAD tumors. Although recent studies have also demonstrated distinct epidemiologic and genomic differences between high and low/intermediate grade tumors^53^, our study is the first to demonstrate in detail the distinct immune differences, as generated by the risk-based tiered classification.

When using risk-based tiered classification, we demonstrate that high-grade tumors appear to represent the morphologic phenotype of an active, heavy smoker, particularly those with greater than 20 pack-years of smoking. Furthermore, our multiplatform analysis reveals that these high-grade tumors have increased TMB, loss of mutations in *TP53*, and increased expression of PD-L1 relative to low/intermediate grade tumors, all of which are independent predictors of response to checkpoint blockade^44,46,47,54^. Finally, we describe the novel finding that tumor microenvironments of high-grade tumors, when classified by risk-based tiered classification, possess a distinct immunogenic phenotype with significant infiltration of cytotoxic CD8^+^ T-cells, increased activated CD4 memory T-cells, and polarization of macrophages toward pro-inflammatory M1. Further, with less shared homology with TAL, infiltrating T-cells are likely more tumor specific and thus more primed for anti-tumor immune response. Altogether, these findings define an early-stage LUAD subset that would be predicted for favorable response to immunotherapy, including standard-of-care checkpoint inhibitors, a finding which is increasingly important in the era of neoadjuvant and adjuvant immunotherapy. It may also define a subset predicted to respond to novel checkpoint inhibitors in development, particularly against LAG-3, for which there is an ongoing clinical trial in NSCLC (NCT03516981) and tumor-infiltrating lymphocyte (TIL) therapy.

Notably, our study does have certain limitations. First, as our cohort of patients underwent surgical resection between 1996 and 2008, pre-surgical staging with PET/CT and endobronchial ultrasound (EBUS) was not performed and could have influenced the difference in clinical stage between groups. Therefore, further study is warranted to determine if our findings would hold true in the current era of pre-surgical staging. Moreover, although we controlled for pathologic stage in our comparison of prognosis, we did not assess for other variables that have been suggested to impact prognosis in early-stage NSCLC such as level of tumor differentiation and pleural or vascular invasion^55,56^. Additionally, for the sake of simplicity, although nearly all LUAD tumors consist of multiple different co-existing patterns of growth and invasion, our analyses evaluated tumors as a whole as opposed to specific analysis of individual growth patterns within a tumor. Therefore, correlations between an individual growth pattern within a tumor and genotypic or immune trends is not possible. However, as stated before, we feel that the design of the risk-based tiered classification compensates for this by accounting for and unmasking contribution of even small high-grade patterns within a tumor.

In conclusion, we suggest that high-grade patterns of growth, of any percentage within a LUAD tumor, possess a distinct immunogenic tumor phenotype that should be considered at the time of surgery and in clinical decision-making regarding adjuvant therapy.

## Supporting information

Legends for Supplementary Figures and Tables

## Data Availability

All data produced in the present study are available upon reasonable request to the authors

## Acknowledgements

This study was supported by the NIH CCSG Award (CA016672 (Institutional Tissue Bank (ITB) and Research Histology Core Laboratory (RHCL)) and the Translational Molecular Pathology-Immunoprofiling Moonshot Platform (TMP-IL) at the Department Translational Molecular Pathology, the University of Texas MD Anderson Cancer Center; the Lung Specialized Programs of Research Excellence grant 5 (P50 CA070907).

AR is supported by the Exon20 Group, Rexanna’s Foundation for Fighting Lung Cancer, the Happy Lungs Project, the Waun Ki Hong Lung Cancer Research Fund, MD Anderson’s Lung Cancer Moon Shot, the Petrin Fund, the University Cancer Foundation via the Institutional Research Grant program at the University of Texas MD Anderson Cancer Center, and the Cancer Prevention and Research Institute of Texas (CPRIT - RP210137). All other authors declare no disclosures.

## Author contributions

N.A., M.L.F., and A.R. wrote the manuscript. N.A., J.Z., J.F., and A.R. designed the study. A.R., C.B., X. H., H.K., R.C., W-C.L., Jianh.Z, P.S., I.W., P.A.F generated and analyzed the data. N.A., A.R., M.L.F, E.M.B., J.Z., and J.F. interpreted the data. JieZ, J.L., and P.S. performed statistical analysis. A.V., B.S., S.S., C.M., N.K, J.H, D.G. J.F., E.R.P, A.W. and I.W. obtained clinical samples and data. J.F., E.R.P, I.W. and A.W. performed pathologic analysis of all samples. JZ, JF, and AR jointly supervised the study. All authors reviewed and edited the manuscript.

