## Supplementary figures and images for "Immunogenomic profiling of lung adenocarcinoma reveals high-grade growth patterns are associated with an immunogenic tumor microenvironment"

### Supplementary Figure 1

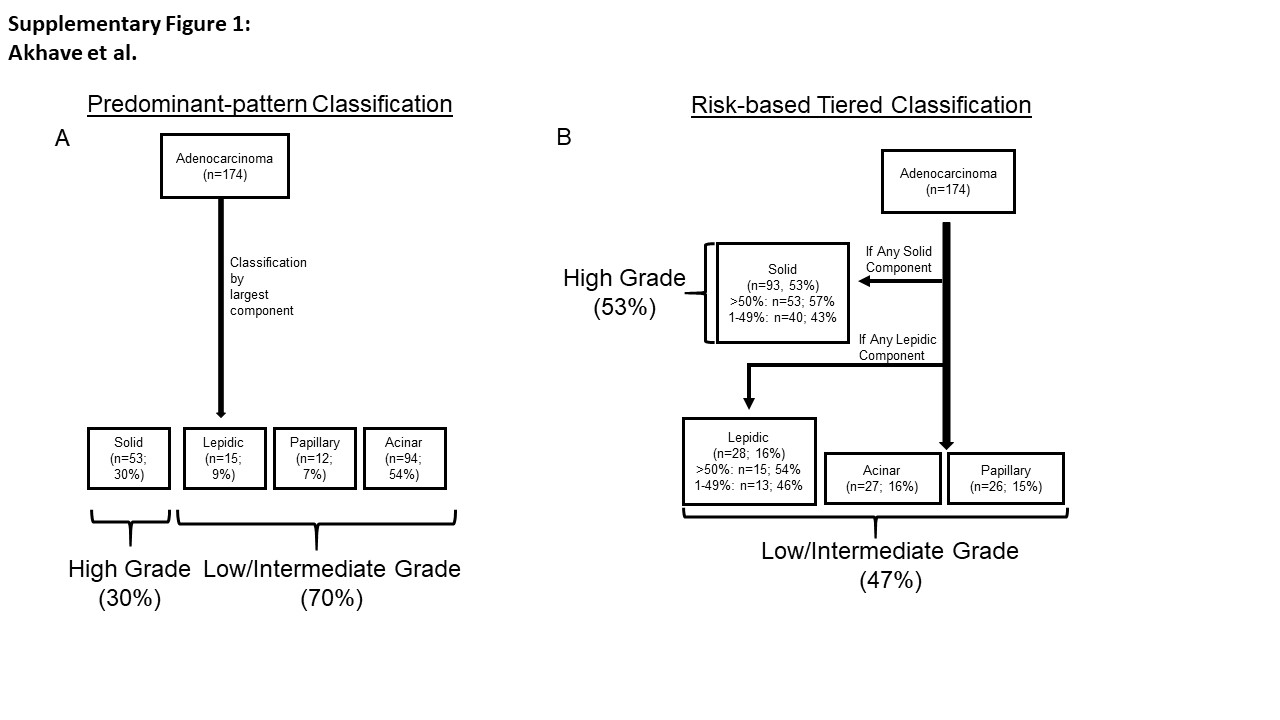

### Supplementary Figure 2

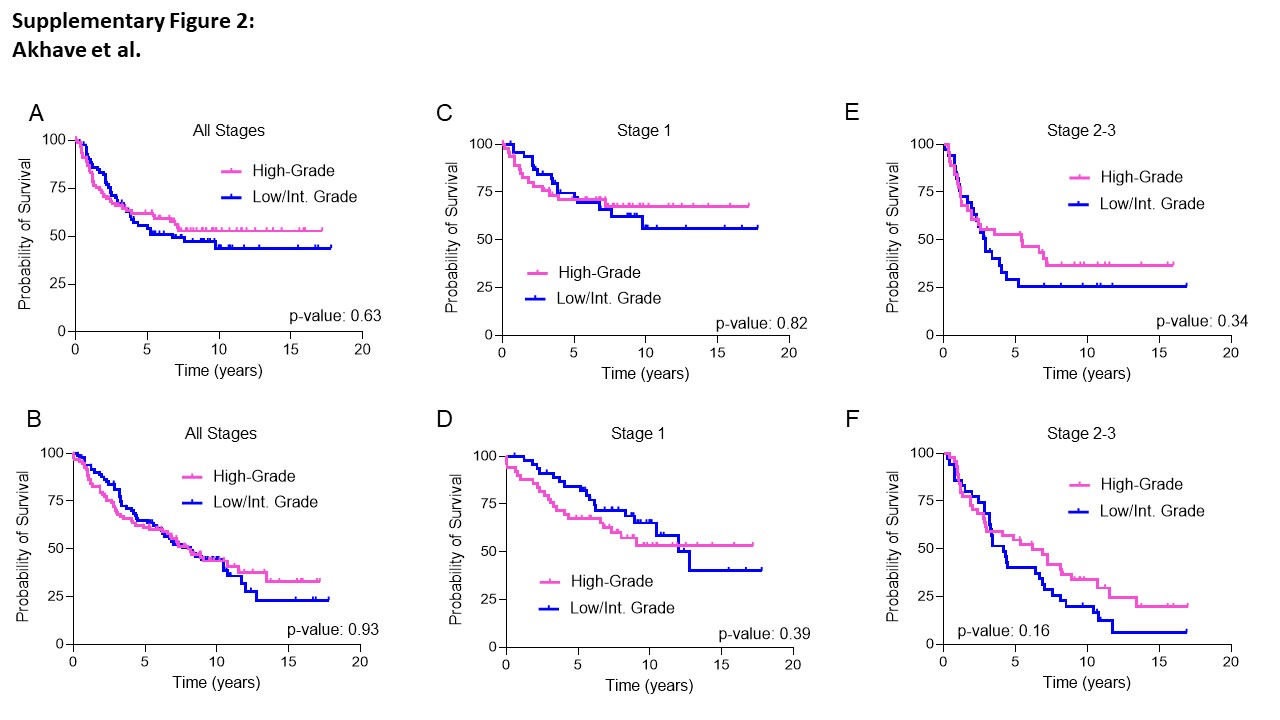

### Supplementary Figure 3

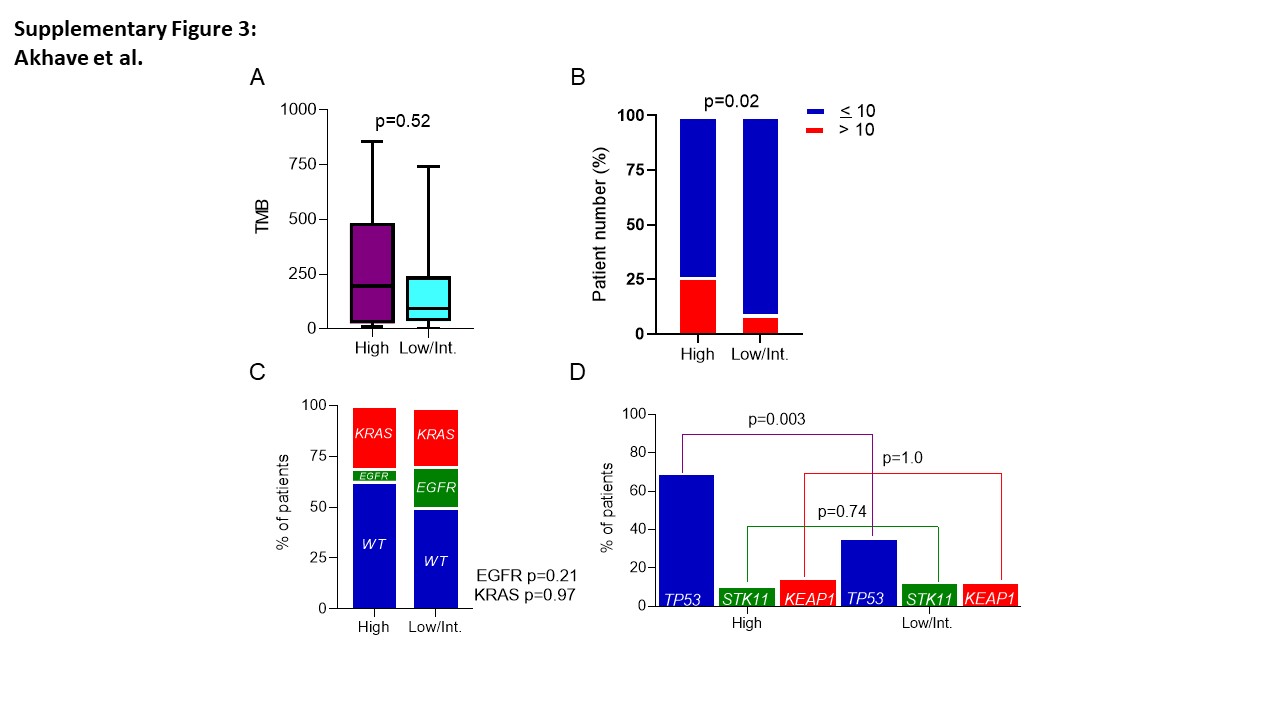

### Supplementary Figure 4

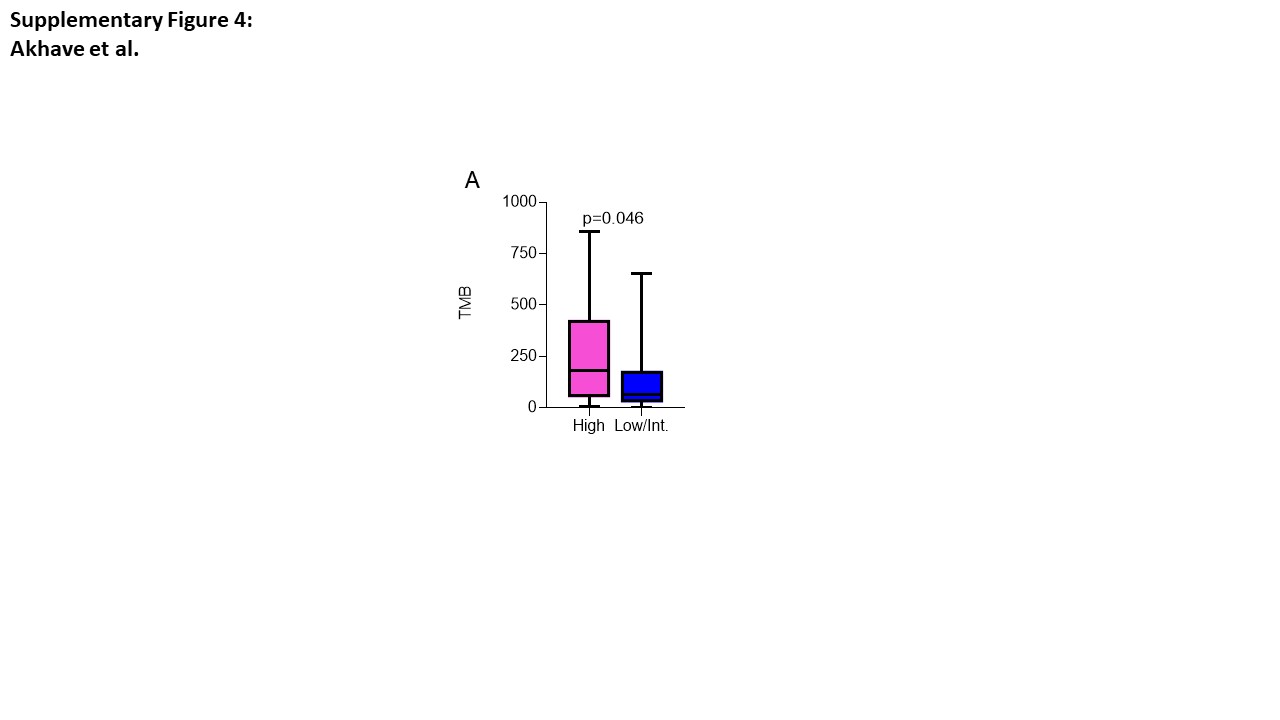

### Supplementary Figure 5

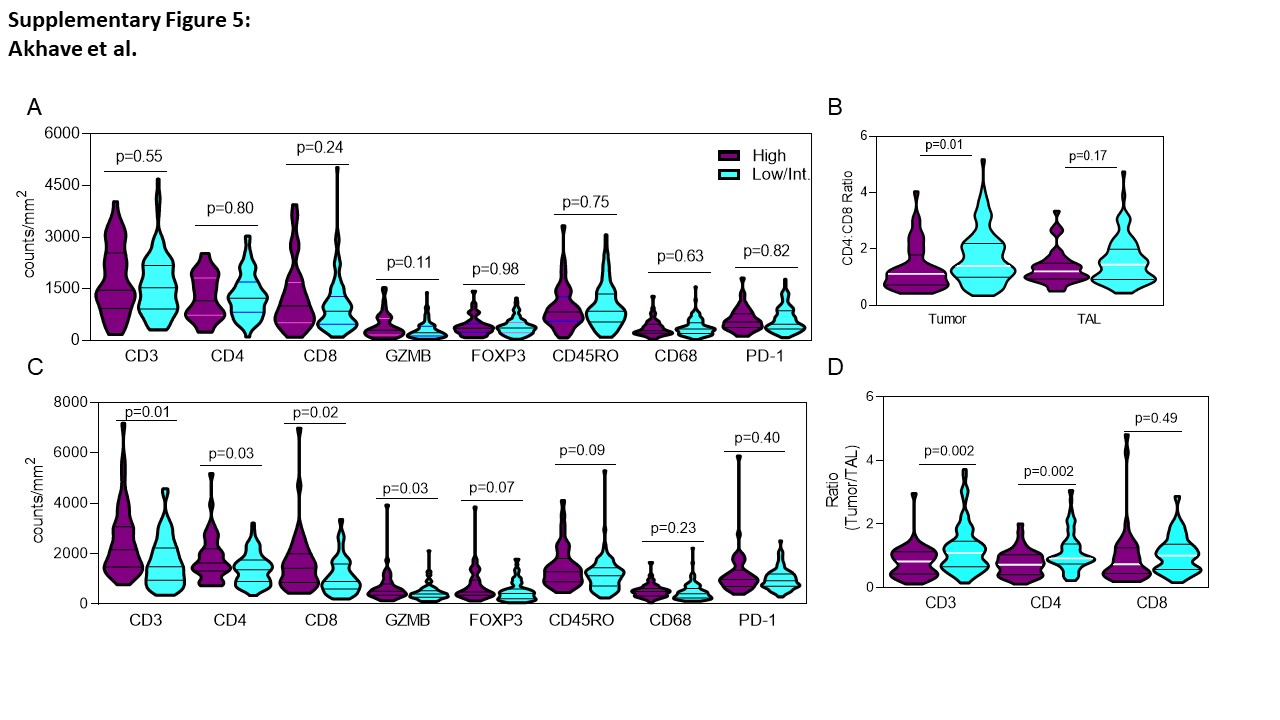

### Supplementary Figure 6

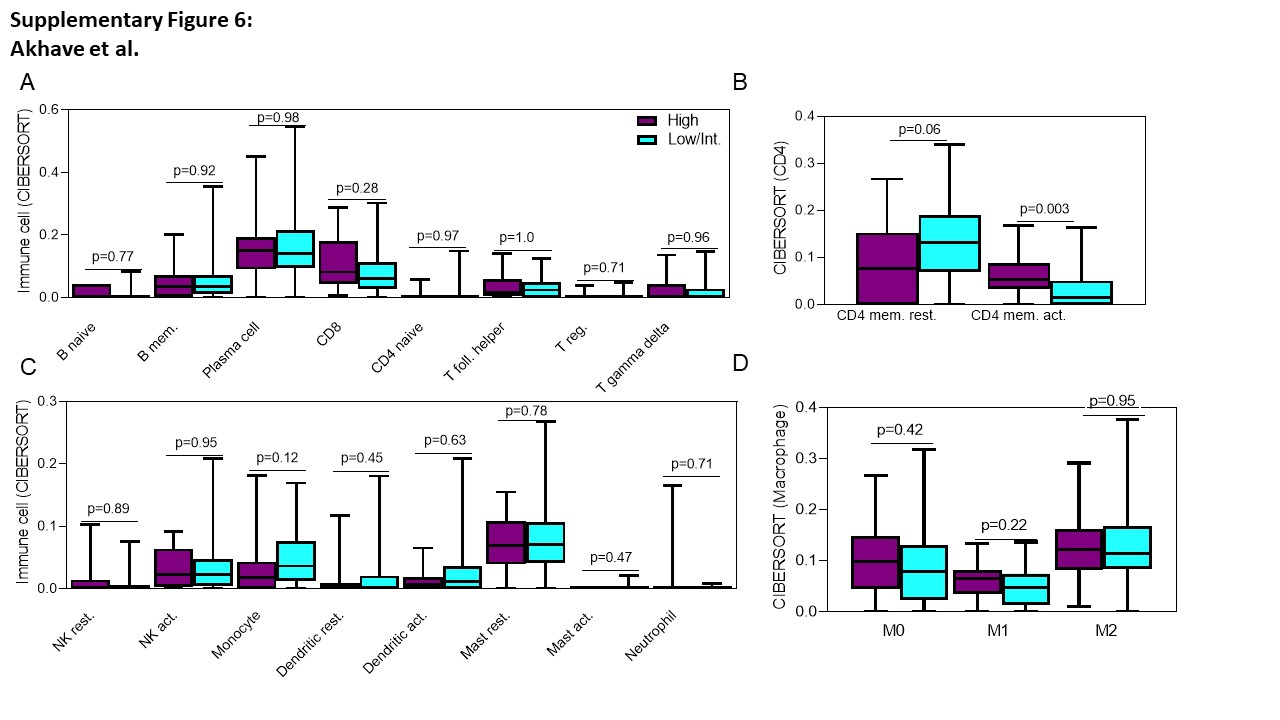

### Supplementary Figure 7

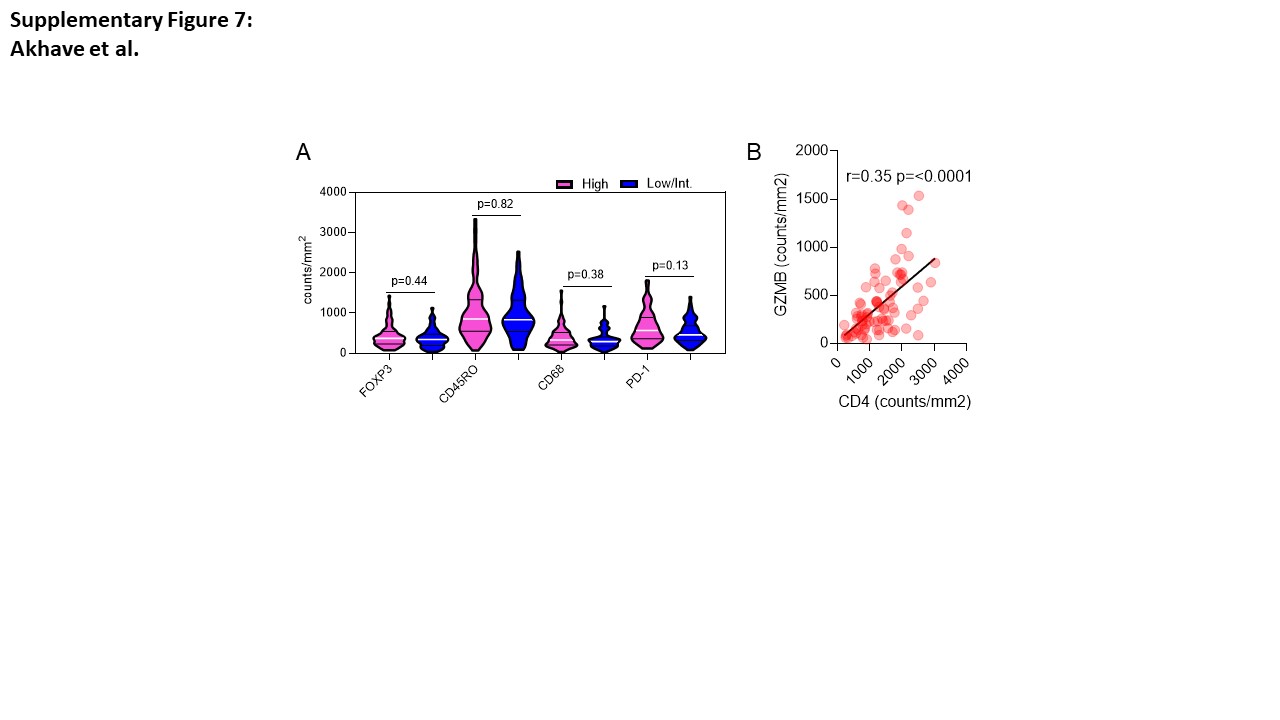

### Supplementary Figure 8

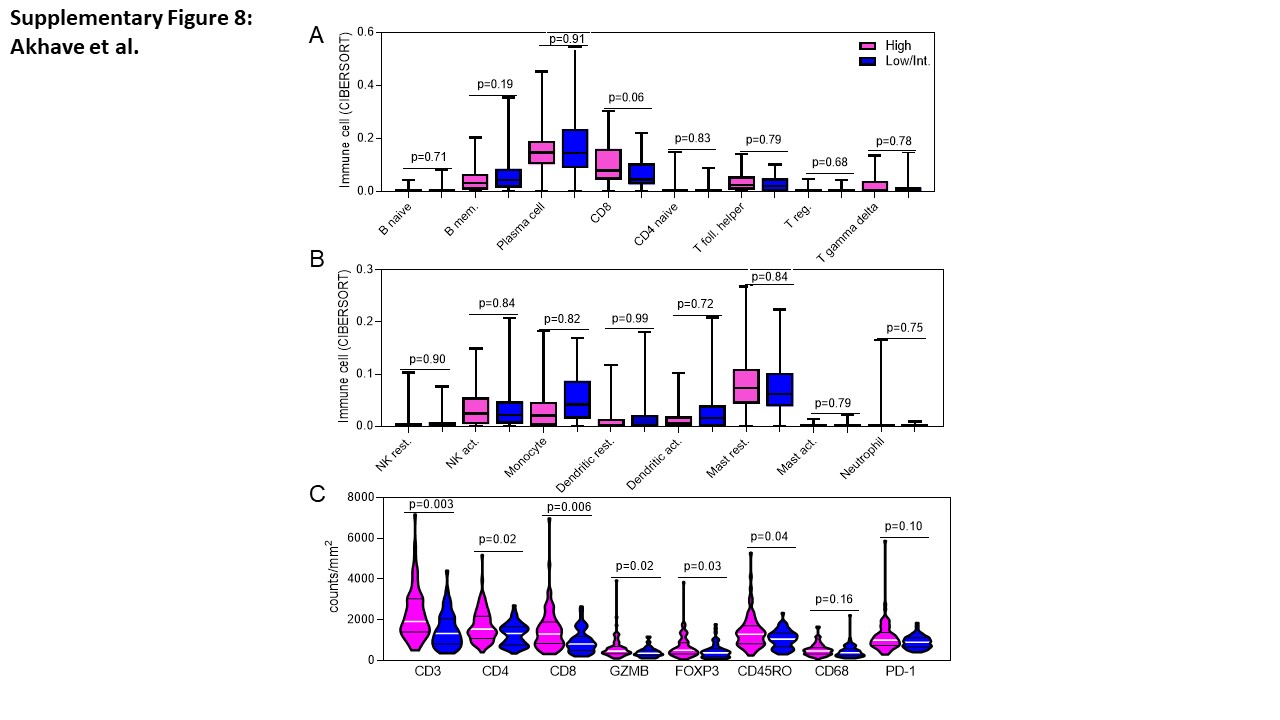

### Supplementary Figure 9

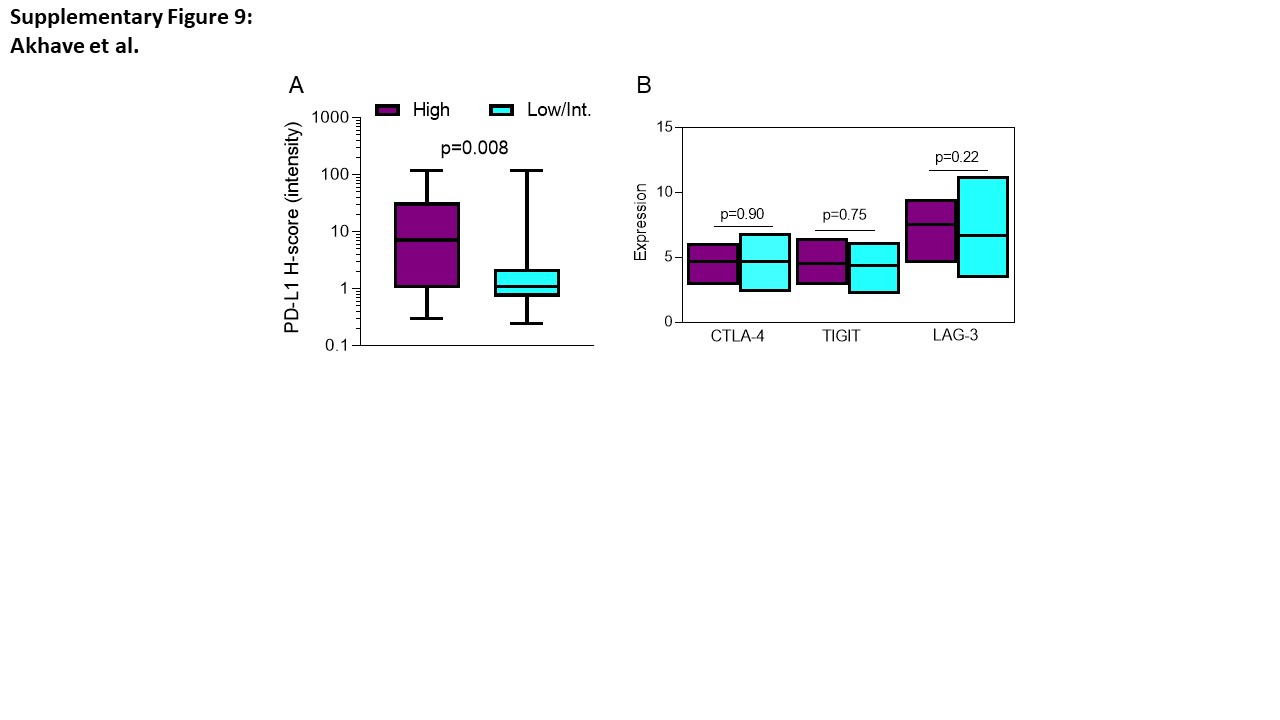

### Supplementary Figure 10

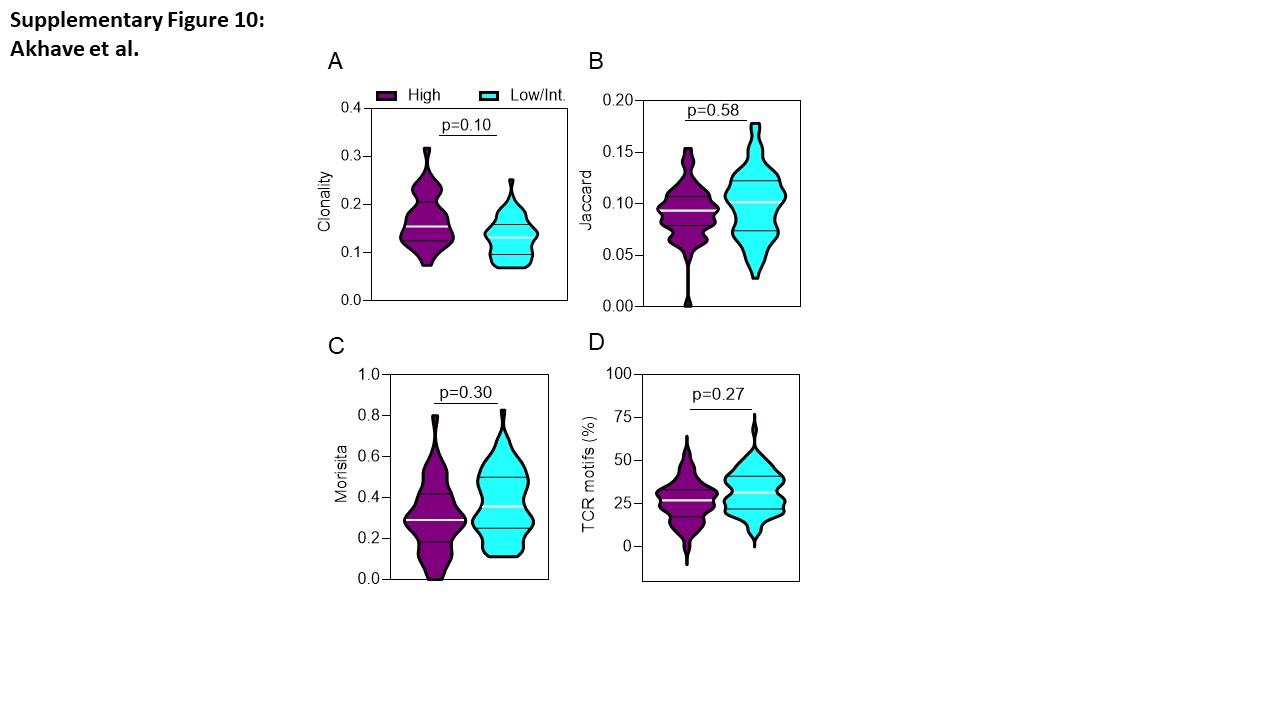

### Supplementary Figure 11

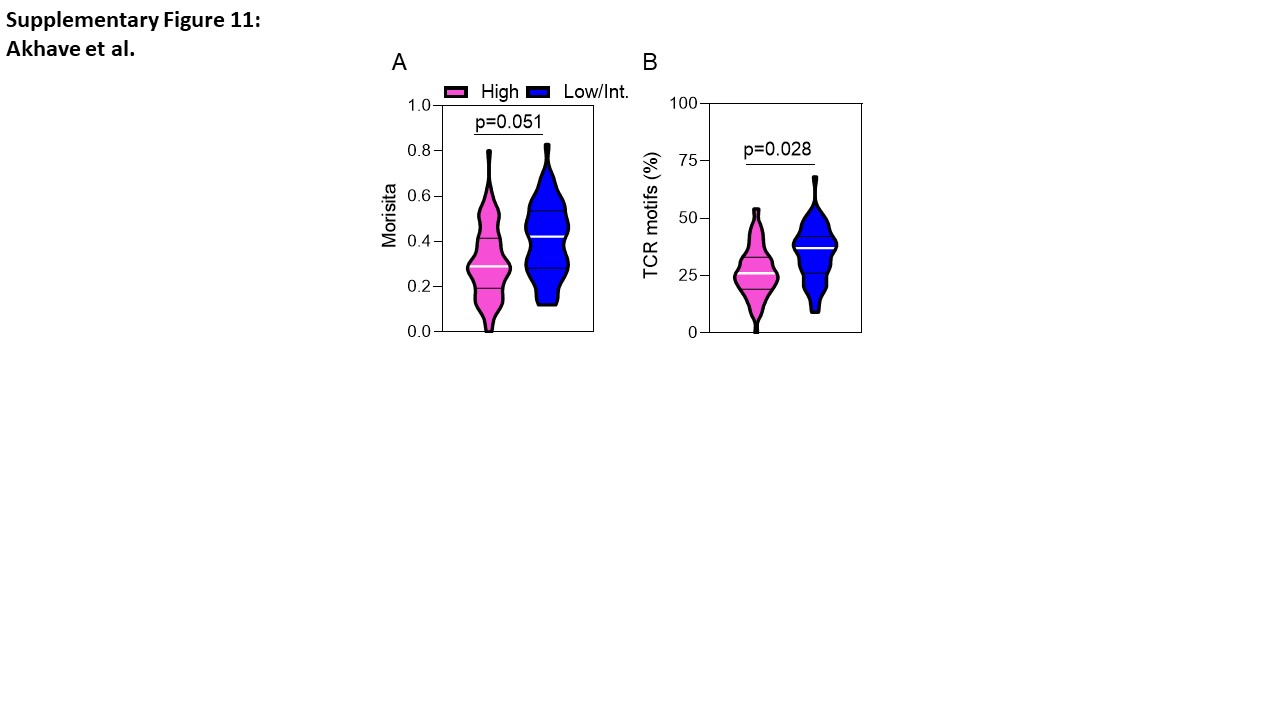

### Supplementary Table 1

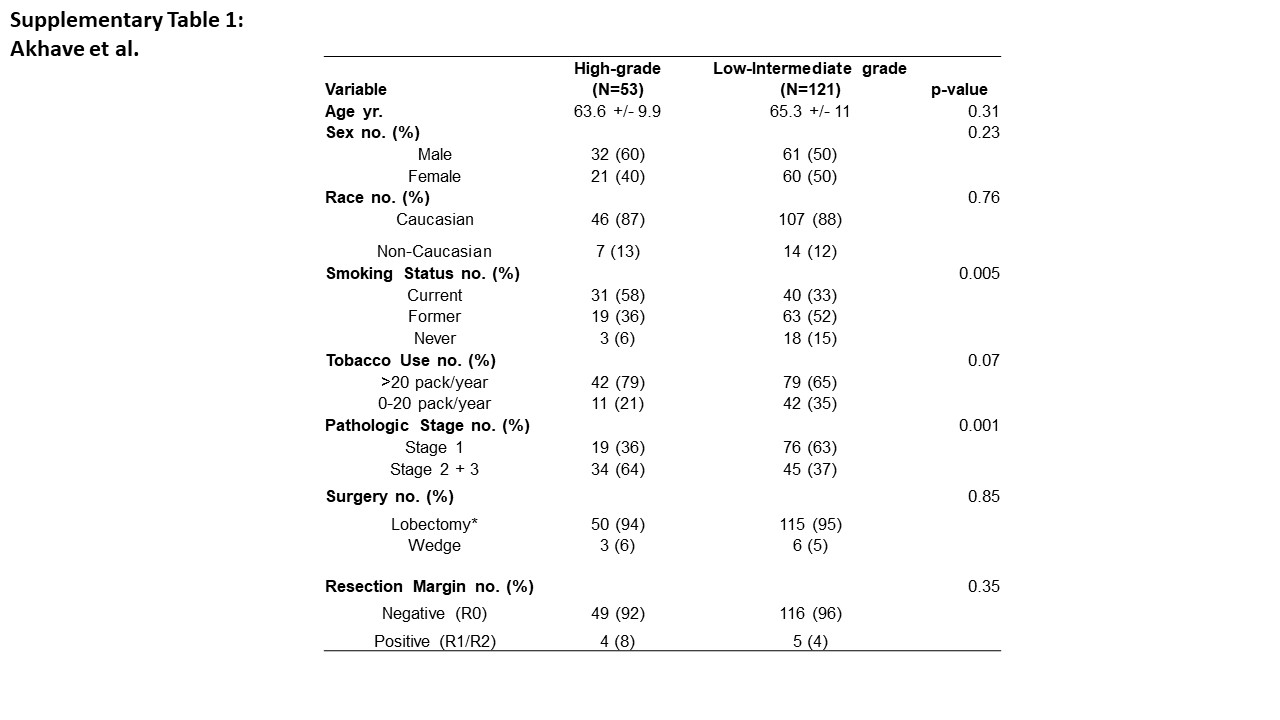

### Supplementary Table 2

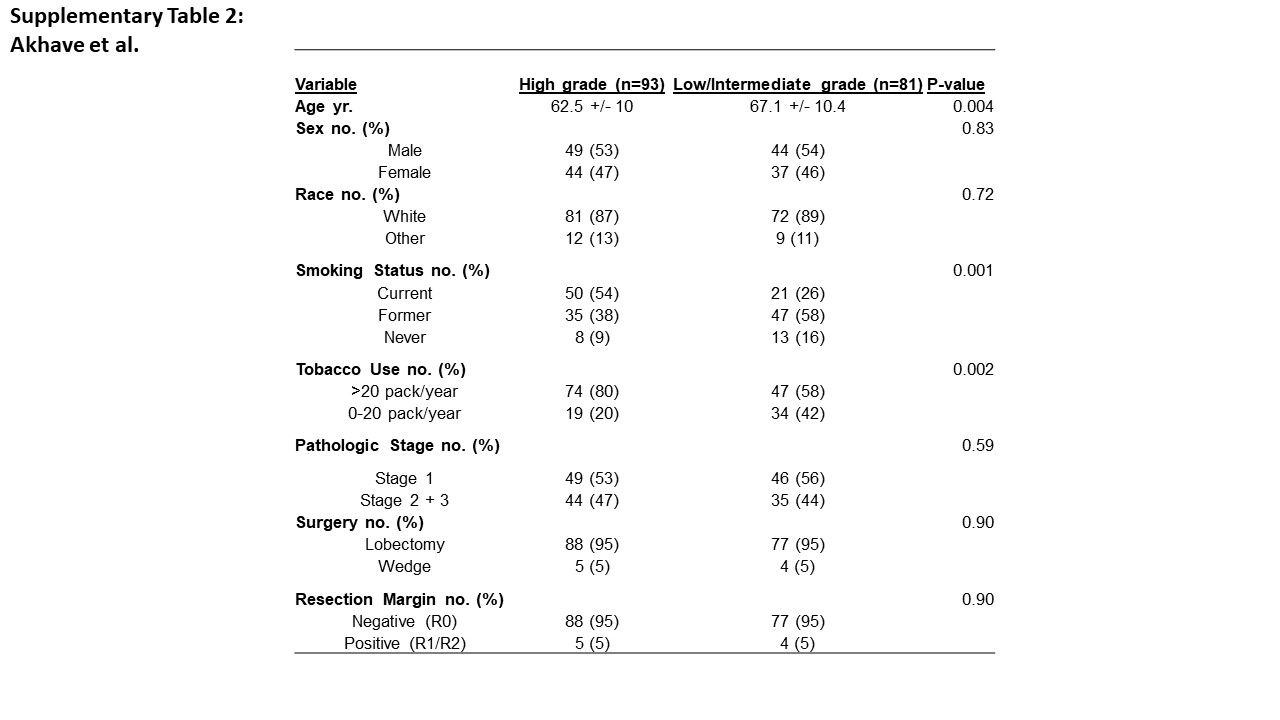
