## Supplementary material for "Immunogenomic profiling of lung adenocarcinoma reveals high-grade growth patterns are associated with an immunogenic tumor microenvironment": Legends for Supplementary Figures and Tables

**Supplementary Table Legends:**

**Supplementary Table 1:** Basic characteristics using predominant-pattern classification method

*Lobectomy also includes those with pneumonectomy

**Supplementary Table 2:** Basic characteristics using risk-based tiered classification method

*Lobectomy also includes those with pneumonectomy

**Supplementary Figure Legends:**

**Supplementary Figure 1:** A. Predominant-pattern (IASLC-ATS-ERS) Classification System. B. Risk-based Tiered Classification System

**Supplementary Figure 2 Risk-based tiered classification does not identify tumors with worse relapse-free and overall survival.** Association between High (violet) and Low/Intermediate (blue) grade tumors and A. relapse-free survival (n=174) and B. overall survival (n=174) independent of stage. C. Relapse-free survival (n=95) and D. overall survival (n=95) when restricted to Stage 1 tumors. E. Relapse-free survival (n=79) and F. overall survival (n=74) when restricted to Stage 2 and 3 tumors.

**Supplementary Figure 3 Predominant-pattern classification identifies high-grade LUAD pattern as associated with TMB (>10 Mut/Mb) and mutations in *TP53***. Relationship between Pattern of growth and A. TMB (by total NSEM) (n=101) and B. TMB (> or < 10Mut/Mb) (n=101). Relationship of Pattern of growth and D. KRAS/EGFR driver mutations (n=162) and TP53/STK11/KEAP1 tumor suppressor genes (n=91)

**Supplementary Figure 4 Risk-based tiered classification identifies high-grade LUAD pattern as associated with TMB**. A. Relationship of Pattern of growth and TMB (n=91)

**Supplementary Figure 5 Predominant-pattern classification identifies high-grade LUAD pattern as associated with distinct TME with increased immune infiltration and pooling in TAL.** A. Relationship of Pattern of growth and intra-tumoral T-cell populations (n=146). B. Relationship of Pattern of growth and CD4:CD8 ratio (n=146). C. Relationship of Pattern of growth and T-cell populations in TAL (n=129). D. Relationship of Pattern of growth and Ratio of CD3, CD4, CD8 T-cell populations between Tumor and TAL (n=129)

**Supplementary Figure 6 Predominant-pattern classification identifies high-grade LUAD pattern as associated with CD4 T-cell activation.** Relationship of Pattern of growth and contents of immune cells by CIBERTSORT. A. B & T-cells (n=139). B. CD4 memory T-cells (n=139). C. NK, Monocyte, Dendritic, Mast, and Neutrophils (n=139). D. Macrophages (n=139)

**Supplementary Figure 7: Risk-based tiered classification identifies high-grade LUAD pattern as associated with distinct immune infiltrated TME** A. Relationship of Pattern of growth and intra-tumoral T-cell populations. B. Among high-grade tumors, relationship of CD4 and GMZB density (n=81).

**Supplementary Figure 8: Risk-based tiered classification identifies high-grade LUAD pattern as associated with distinct immune infiltrated TME** CIBERSORT of A. Adaptive immune cell (n=139) and B. Innate Immune cell populations (n=139) C. Relationship of Pattern of growth and TAL T-cell populations.

**Supplementary Figure 9: Predominant pattern classification identifies high-grade LUAD pattern as associated increased expression of PD-L1 but not other immune checkpoints.** Relationship of pattern of growth and A. PD-L1 H-score by IHC (n=146) and B. CTLA4, TIGIT, and LAG3 by GEP (n=101)

**Supplementary Figure 10: Predominant pattern classification does not demonstrate differences in T-cell clonality or T-cell repertoire homology.** Relationship of Pattern of growth and A. T-cell clonality (n=104), B. Jaccard index (n=130), C. Morisita index (n=130), and D. shared Top 100 T-cell clones in tumor relative to Top 100 T-cell clones in TAL (n=130)

**Supplementary Figure 11: Risk-based tiered classification identifies high-grade LUAD pattern of growth as associated with decreased T-cell repertoire homology between tumor and TAL** Relationship of Pattern of growth and A. Morisita index (n=130) and B. Shared Top 100 T-cell clones in tumor relative to Top 100 T-cell clones in TAL (n=130)
